# The longitudinal kinetics of antibodies in COVID-19 recovered patients over 14 months

**DOI:** 10.1101/2021.09.16.21263693

**Authors:** T. Eyran, A. Vaisman-Mentesh, Y. Dror, L. Aizik, A. Kigel, S. Rosenstein, Y. Bahar, D. Taussig, R. Tur-Kaspa, T. Kournos, D. Markovitch, D. Dicker, Y. Wine

## Abstract

Here, we describe the longitudinal kinetics of the serological response in COVID-19 recovered patients over the period of 14 months. The antibody kinetics in a cohort of 200 recovered patients with 89 follow up samples at 2-4 visits reveal that RBD-specific antibodies decay over the period of 14 month following the onset of symptoms. The decay rate is associated with the robustness of the response thus, recovered patients that exhibit elevated antibody levels at the first visit, experience faster decay. We further explored the longitudinal kinetics differences between recovered patients and naïve BNT162b2 vaccinees. We found a significantly faster decay in naïve vaccinees compared to recovered patients suggesting that the serological memory following natural infection is more robust compared to vaccination. Our data highlights the differences between serological memory induced by natural infection vs. vaccination, facilitating the decision making in Israel regarding the 3^rd^ dose vaccination.

## Introduction

The first patients detected positive to coronavirus disease 2019 (COVID-19) were identified in Wuhan, China ^1^. These patients were found to be infected by severe acute respiratory syndrome coronavirus 2 (SARS-CoV-2) leading to the declaration of the World Health Organization (WHO) that COVID-19 is a worldwide pandemic ^2^. Rapid response to the outbreak provided important information regarding the virus genome sequence and especially the Spike protein (S protein) and its sub-region, the receptor binding domain (RBD) which is responsible for the binding to the human angiotensin-converting enzyme 2 (hACE2) to mediate virus entry ^3^. Measuring antibody responses to SARS-CoV-2 antigens has been vital for ascertaining past viral exposure, investigating transmission in the community and carrying out serosurveys ^4^. The longitudinal kinetics of the antibody immune response following COVID-19 recovery is essential for evaluating the persistence of serological memory. The S protein is highly immunogenic with the RBD holding the potential to elicit neutralizing antibodies (nAbs) that blocks the interaction with hACE2 ^5^, leading to viral neutralization and, as such, can be used as marker of functional immune responses ^6^. Utilizing RBD as a marker is further supported by the correlation between the RBD-specific (RBD^+^) antibody levels and neutralization capacity ^7,8^.

The kinetics of SARS-CoV-2 antibodies can be evaluated at the acute course of disease and following recovery. The S protein RBD, elicits antibodies starting as early as 5-15 days following symptoms ^9,10^ in which their levels increase along the progression of the disease ^10-13^. However, longitudinal kinetics of viral-specific antibodies in COVID-19 recovered patients is not fully clear and there have been contradicting reports regarding the persistence of anti-SARS-CoV-2 antibodies ^14-19^. Specifically, it was estimated that the half-life of SARS-CoV-2 anti-spike antibodies can reach > 7 month ^20^ and some showing persistence over 10 months ^21-23^. A recent study that examined long-lived plasma cells (LLPC) in the bone marrow estimated that SARS-CoV-2 infection induces a robust antigen-specific, long-lived humoral immune response in humans ^24^. The majority of studies that examined the antibody kinetics following COVID-19, were cross-sectional however, antibody response kinetics in patients who have recovered from COVID-19 vary greatly, thus evaluation of immune longevity can only be accurately determined at the individual level. Moreover, most follow up longitudinal studies were limited in terms of duration following onset of symptoms thus, the information regarding the persistence of the serological memory is time restricted.

We conducted a longitudinal study on samples obtained from 200 patients who had recovered from COVID-19 up to 450 days following onset of symptom by recording the changes in RBD^+^ antibody level. Follow up samples from 89 COVID-19 recovered patients were collected across 4 visits (V1-V4) with an interval of approximately 90 days. Immunoglobulin G (IgG), IgM and IgA were measured and kinetic parameters were calculated. First, we found that the RBD^+^ IgG/M/A levels wane over the period of 14 months with IgA showing the fastest decay rate followed by IgM and IgG. We further analyzed the decay kinetics by stratifying the antibody levels to quartiles according to the antibody levels detected at the first visit (V1). Here we found that the decay was significantly faster in patients that exhibited antibody levels at V1 that were designated to the highest quartile.

Additionally, we analyzed the kinetics of the antibody levels in a cohort of COVID-19 recovered patients that received one BNT162b2 mRNA vaccine dose (n=20). We show that the decay progression was halted and the vaccine induces a robust response giving a rise to antibodies that exceeds the levels observed prior to the decay. Lastly, samples collected from naïve vaccinees (two doses of BNT162b2 mRNA-vaccine), were used to compare their antibody kinetics with the kinetics of COVID-19 recovered patients. We found that naïve vaccinees exhibited a significantly faster antibody decay compared to the recovered cohort. Overall, our data provides new insights regarding the longitudinal kinetics of COVID-19 recovered patients that may help decision making regarding vaccination regimes.

## Results

### Cohort establishment and antibody measurement

To investigate the longitudinal kinetics and persistence of SARS-CoV-2 specific antibodies, we established a cohort of 200 COVID-19 patients who recovered from SARS-CoV-2 infection. The cohort included individuals that were positive to SARS-CoV-2 as determined by qPCR and were admitted to the HaSharon hospital, Rabin medical center, Israel. Days following onset of symptoms (DFS) was derived from self-reported symptom onset. COVID-19 recovered patients enrolled cohort consists of: median age of 53 years (range: 20 to 81 years), gender distribution of males and females was balanced, and disease severity included mild (83%) and moderate/severe (17%) cases. Blood samples were collected from all participants at visit 1 (V1, mean 90 DFS) with follow up collections at visit 2 (V2) from 55.5% participants (mean 184 DFS), visit 3 (V3) from 32.2% participants (mean 298 DFS) and visit 4 (V4) from 12.2% participants (mean 406 DFS) (**Figure 1a-b; Table S1**).

**Fig. 1.**
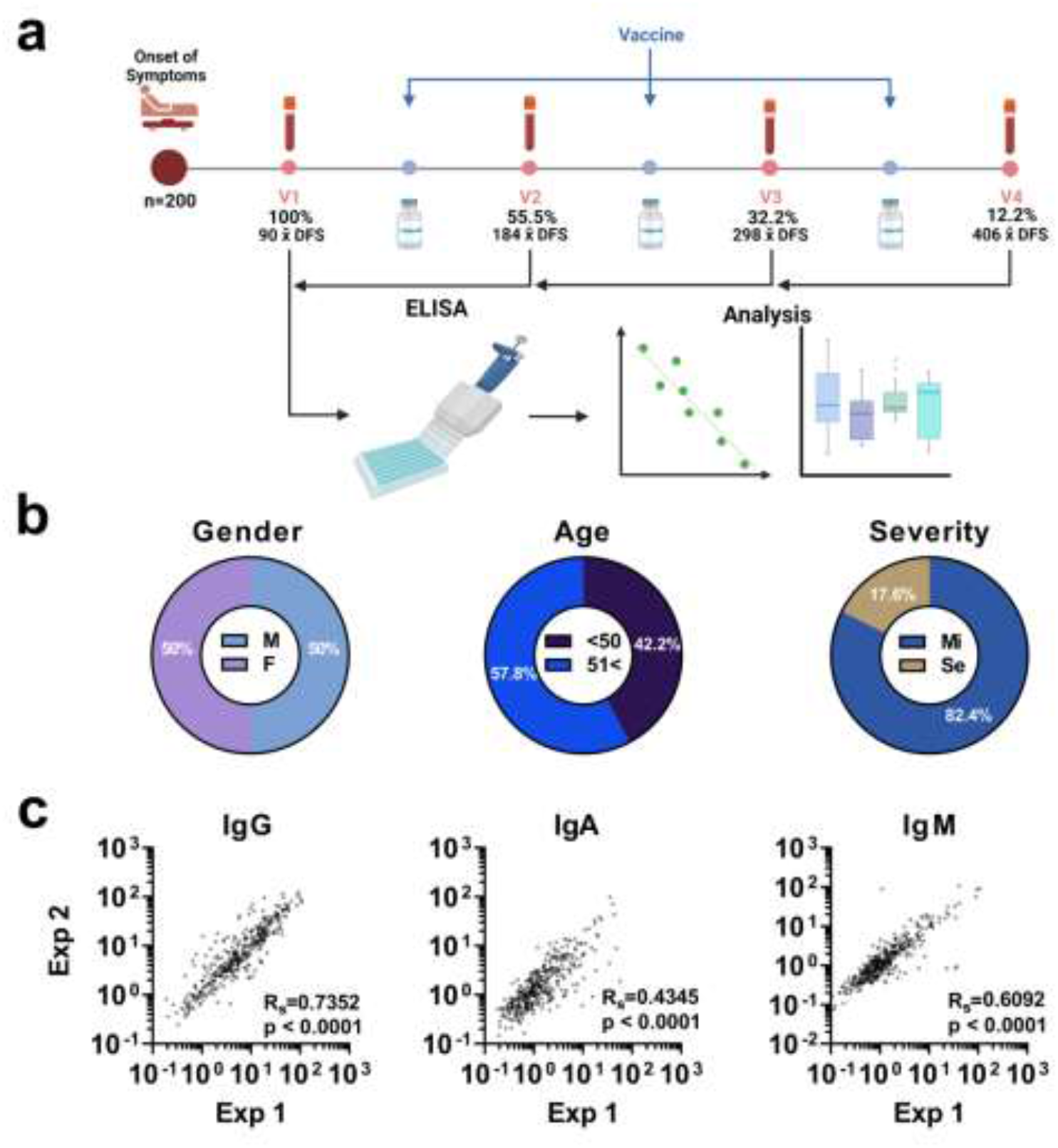
Experimental design, cohort characteristics, and reproducibility of antibody level measurements. **a**) 200 COVID-19 recovered patients were recruited to the study at V1 with 89 patients donating blood samples at subsequent visits (V2-V4) at intervals of approximately 3 months between visits. A subset of the recruited patients, received one dose of mRNA-vaccine at various time points DFS. Blood samples were fractionated for the isolation of plasma samples that were used to determine the RBD^+^ antibody levels and for the analysis of the longitudinal kinetics. **b**) The cohort of COVID-19 recovered patients included a balanced male/female population and age distribution below and above 50 years. The majority of the recovered COVID-19 patients exhibited mild symptoms. **c**) RBD^+^ measurements were carried out in duplicates and each experiment was repeated twice (designated Exp1 and Exp2). Spearman’s rank correlation (RS) was used to determine the reproducibility of the experiments. p values < 0.05 were considered significant and Spearman’s rank coefficient is indicated (R_S_)

The sub-cohort of COVID-19 recovered patients (24%) were vaccinated with the BNT162b2 COVID-19 mRNA vaccine within a time window of approximately 222 DFS.

Additionally, we included a prospective cohort of 17 naïve individuals that received two doses of the BNT162b2 mRNA-vaccine. Samples from this cohort were collected at four visits following mRNA-vaccine 2^nd^ dose as follows (median, days following vaccination -DFV): 8, 35, 91, and 182 DFV.

RBD^+^ IgG, IgM and IgA levels were measured by semi-quantitative ELISA and antibody levels were normalized by calculating the ratio of the signal over the mean signal obtained in the negative controls used in each microwell plate (signal over background).

Each sample was measured in duplicates and each experiment was repeated independently twice showing high reproducibility (**Figure 1c**). In total, 200 samples were collected for V1 with another 145 subsequent samples for V2-V4 to determine the kinetics of RBD^+^ antibodies within 14 months following symptoms.

### Antibody levels in association with age, gender, and disease severity

To test whether the robustness of the antibody response following SARS-CoV-2 infection is associated with patients’ age, gender and the severity of the COVID-19 disease, RBD^+^ antibody levels at V1 were stratified according to the above-mentioned parameters. We found that the IgG and IgA levels significantly differ between male and females and IgM was not significantly different between gender groups (**Figure 2a**).

**Fig. 2.**
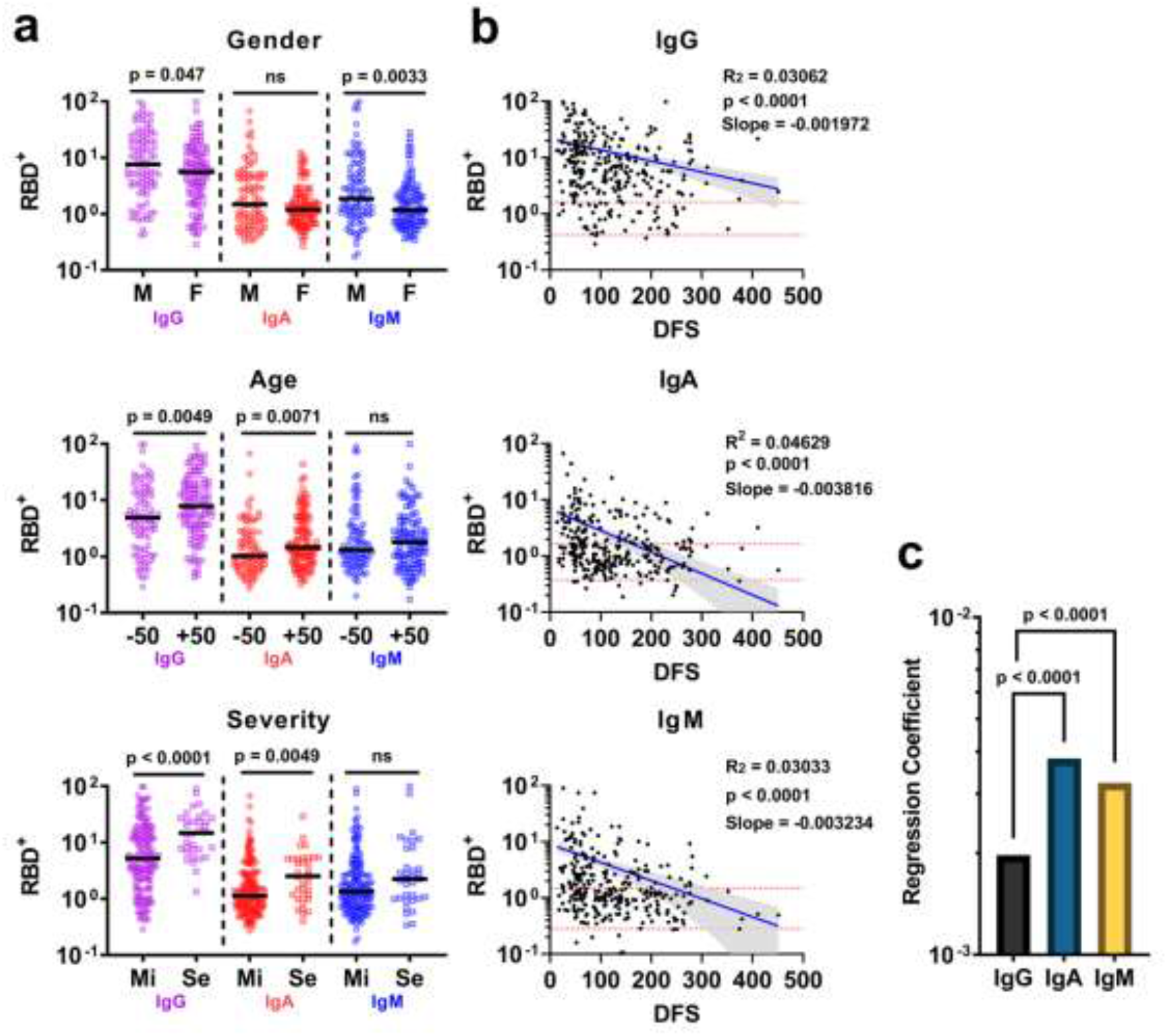
SARS-CoV-2 RBD^+^ antibody levels over the period of 14 months. **a**) grouped scatter plots of RBD^+^ IgG/A/M levels in COVID-19 recovered patients at V1, by gender, age and COVID-19 severity. Y-axis - log_10_ values of the ratio between the signal obtained over the mean signal for the negative controls in each microwell plate. Mean values are indicated by solid black lines. P values were determined with an unpaired, two-sided Mann-Whitney U-test; p < 0.05 was considered statistically significant **b**) scatter plot of RBD^+^ antibody kinetics. X-axis represents days following symptoms (DFS). A non-linear regression model was used to determine the regression coefficient. Negative regression coefficients (i.e., slope) were determined for IgG, IgA, and IgM, indicating that all isotypes are decaying over the period of 14 months **c**) bar plot comparing the regression coefficient obtained for IgG, IgA and IgM. For all plots, p values, R^2^ and slopes are indicated. p values < 0.05 were considered significant.

The cohorts were also divided by age groups with 50 years as the differentiating pivot. Here we found the IgG and IgA levels were significantly elevated in the >50 group compared to <50 groups while IgM did not significantly differ between age groups. The severity during the active disease phase was found to be associated with higher IgG and IgA levels at V1 while IgM was not found to be associated with severity (**Figure 2a**).

### Antibody levels decay over the period of 14 months

All sera samples from COVID-19 recovered patients that were collected during the study (n=345) were used to evaluate the persistence of the RBD^+^ antibodies over the period of 14 months. We examined the association between DFS and the RBD^+^ IgG/A/M levels by applying a regression model and the extent of change over time was determined by the regression coefficient (i.e., the slope of the regression). For all antibody isotypes, the RBD^+^ levels were found to decay over the period of 14 months (**Figure 2b**). The negative values of the regression coefficient indicated that all isotypes exhibit a decline trend, and the regression coefficients differ significantly between isotypes, where IgA showed significantly faster decay followed by IgM and IgG (**Figure 2c**). As a result of the antibody decay, 18.4%, 61.81%, and 54.86% of serum samples (IgG, IgA, and IgM respectively) were found to reach the antibody range determined for negative control samples.

### The temporal decay of antibodies is associated with the robustness of the early antibody response

The longitudinal kinetics of RBD^+^ antibodies at the individual level was determined by measuring the RBD^+^ antibody levels in follow up samples from 89 recovered COVID-19 patients. We found a marked decay in all follow up patients. The fraction of patients that exhibited antibody levels at V1 within the range of the negative control was 14%, 49% and 40% for IgG, IgA, and IgM respectively. The fraction of patients that exhibited antibody levels at their last visit within the range of the negative control was 29%, 70% and 73% for IgG, IgA, and IgM respectively (**Figure 3a**). Next, we stratified the RBD^+^ antibody levels to quartiles by bootstrapping the obtained values from serum samples of the extended cohort (n=345), and quartile thresholds were set according to the mean values derived from the bootstrap iterations (**Figure S1**). The quartile threshold values were then used to designate each measured antibody level at V1 to its respective quartile range (i.e., from low to high Q1-Q4 respectively, **Figure S2**). Follow up samples from the corresponding patients were used to determine the regression coefficient for each quartile group. For example, patients that exhibited RBD^+^ antibody levels at V1 above the Q4 threshold, were used to determine the regression coefficient based on these patients’ follow up samples obtained in the subsequent visits (**Figure 3b**). We found that for all isotypes, patients that were designated to quartile Q4, Q3 and Q2, exhibit negative regression coefficient values indicating that the antibody levels are declining over time. The antibody levels for patients that were designated to quartile Q1, did not show a significant decrease in antibody levels over time. Additionally, the rate of decay as determined by the regression coefficient, suggests that the decay rate is significantly faster for those patients that the antibody levels at V1 was designated to Q4 (**Figure 3c**). This decay was further observed when we examined the regression coefficient for groups Q3 and Q2. For group Q1 there was no significant decay in antibody levels for all isotypes.

**Fig. 3.**
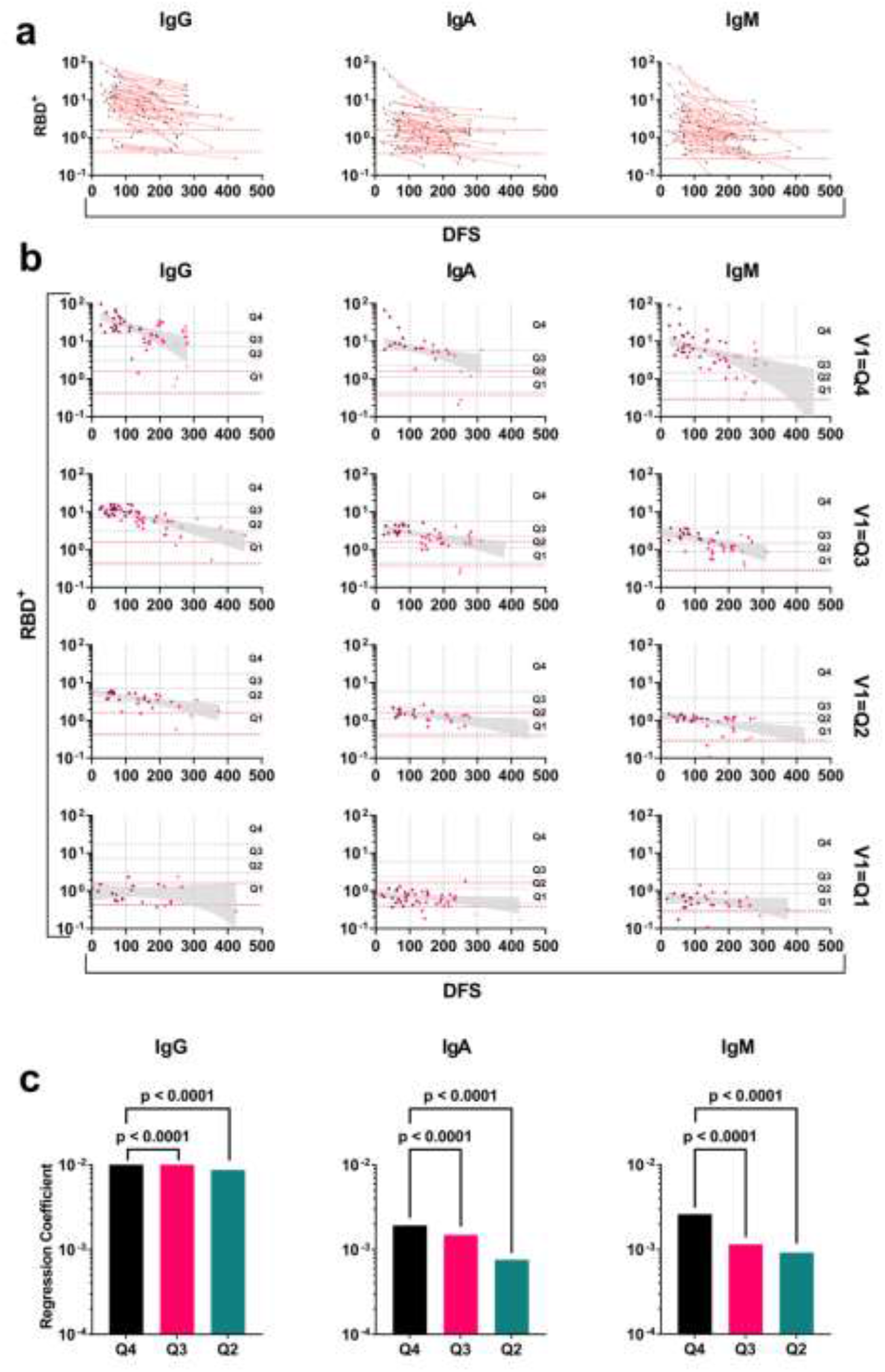
Longitudinal kinetics of RBD^+^ antibodies in COVID-19 recovered patients. **a)** line graph of RBD^+^ antibody levels in COVID-19 recovered patients with follow up samples obtained at 2-4 visits. Y-axis in log_10_ and X-Axis, days following symptoms (DFS). Dashed red lines represent the maximum and minimum range of the antibody levels as measured in negative control samples (n=27). **b)** scatter plots of RBD^+^ antibody levels stratified by quartiles. The antibody levels at V1 determined the attributed quartile and follow up sample from the same patients were used to determine the regression coefficients (slope). Four quartiles for each isotype were analyzed. Quartiles Q4-Q1 thresholds are indicated by horizontal dashed black lines. Y-axis, log_10_, X-axis days following symptoms (DFS). Dashed red lines indicate the upper and lower range of negative controls.

### Antibodies in COVID-19 recovered patients decay slower than in naïve vaccinees

We were interested to examine if the longitudinal kinetics of antibodies following recovery differs from the kinetics following vaccination. To this end, we measured the antibody levels in samples collected at four visits from 17 naïve individuals who received 2 doses of mRNA-vaccine starting on an average of 8 DFV and follow up samples at an average of 35, 91 and 182 DFV. Based on the quartile stratification approach, the RBD^+^ IgG levels at the average of 8 DFV, in all 17 naïve individuals, was designated to the highest quartile. This was not the case for IgA and IgM that were distributed across quartiles. Furthermore, the IgG/A/M levels in all 17 individuals reached the negative control range at the approximate 180 DFV (Figure 4a). The rate of the antibody decay as determined by the regression coefficients reveal that, as with COVID-19 recovered patients, IgA exhibited the fastest decay followed by IgG and IgM (Figure 4b). As the antibody levels at V1 in naïve vaccinees was designated to Q4 we compared the regression coefficient for IgG as measured in the recovered patient group that was designated to Q4 at V1. We found that the decay rate in COVID-19 recovered patients was significantly slower compared to the decay in naïve vaccinees (Figure 4c-d). Moreover, 100% of the naïve vaccinees exhibited antibody levels lower than the upper limit of the control group. On the other hand, only 5% of the recovered patients reached the negative control range at the 182 DFV time point of naïve vaccinees and 19% at an average of 265 DFS in their V4 samples. Next, we examined the effect of vaccination in a subset of the recovered cohort (n=20). As recovered patients received a vaccine at various DFS, we extrapolated the antibody levels to the day that the patients received the vaccine using the regression formula. The antibody levels at 8 DFV were extrapolated as well by using the regression formulas obtained from naïve vaccinees (Figure 4e). Based on the calculated temporal kinetics we found that approximately 80% of recovered vaccinees experienced an increase in IgG levels reaching the highest quartile (Q4) as opposed to 50% of them being designated to Q4 following recovery (Figure 4f). In addition, we found that the predicted IgG levels at 8 DFV was significantly higher than the predicted IgG levels on the day of vaccination (due to continued decay following recovery) and not significantly higher than the antibody levels at V1 (Figure 4g). Still, the increase in IgG levels 8 DFV in recovered patients was not significantly higher than the antibody levels detected 8 DFV in naïve vaccinees who received 2 doses of the mRNA-vaccine (Figure 4g).

**Fig. 4.**
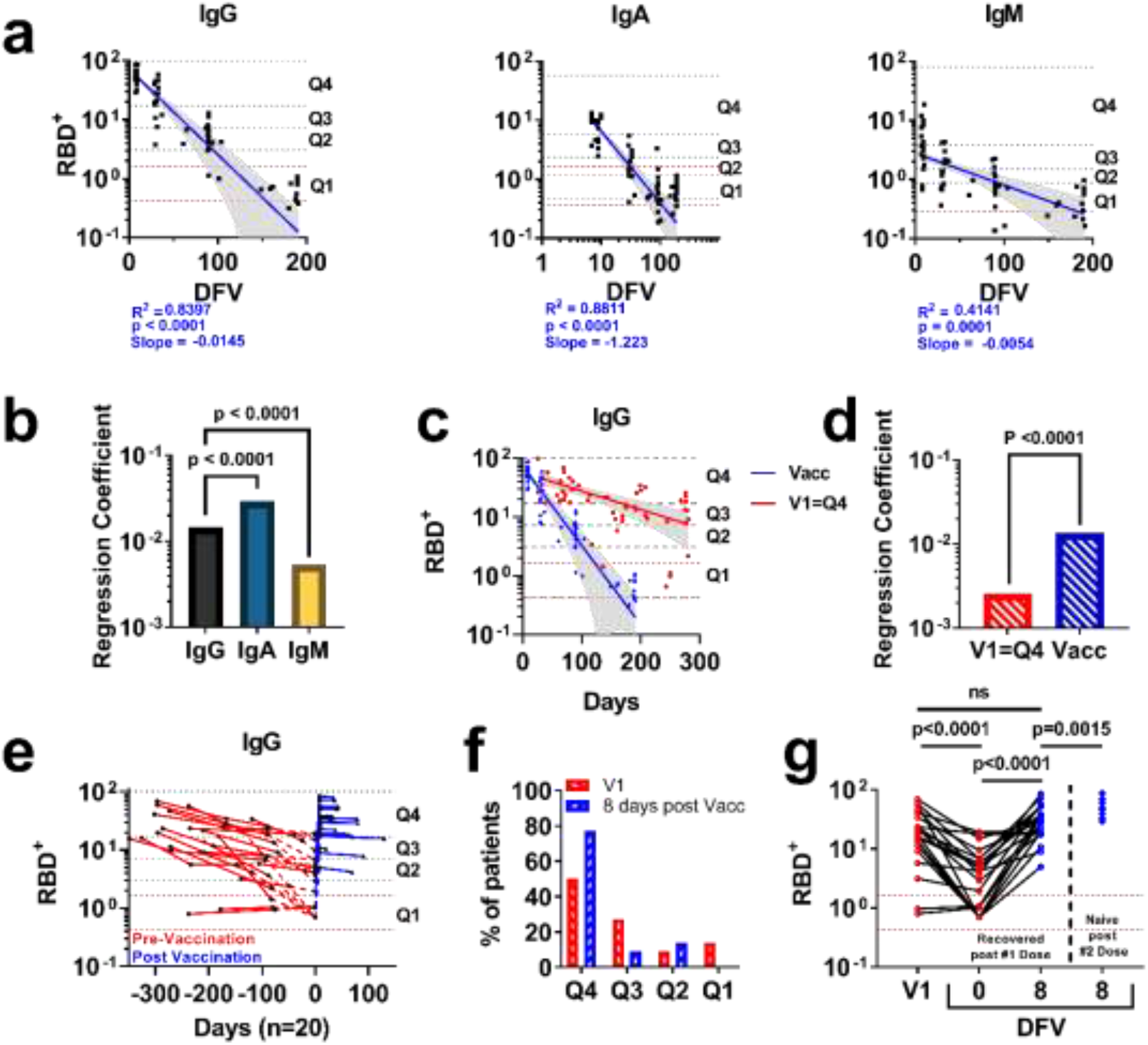
RBD^+^ antibody kinetics differ between recovered patients and naïve vaccinees. **a**) RBD^+^ IgG/A/M kinetics of naïve vaccinees following 2 vaccine doses. Nonlinear regression model was applied, and the regression coefficient was determined. The 95% CI is plotted in grey. Y-axis log_10_ values. X-axis represents days following vaccination (DFV). Dashed red lines indicate the upper and lower limit of negative control samples. Horizonal dashed black lines indicate the quartiles. **b**) Bar plot comparing the regression coefficient obtained for IgG, IgA and IgM. For all plots, p values, R^2^ and slopes are indicated. p values < 0.05 were considered significant. **c**) RBD^+^ IgG/A/M kinetics in naïve vaccinees in comparison with the kinetics in recovered patients whose IgG levels were designated as Q4 at the first visit (V1). Y-axis log_10_. X-axis represents days following symptoms for recovered patients (red) and days following vaccination for naïve vaccinees (blue). **d**) Multiple comparison between the regression coefficients in recovered patients and naïve vaccinees. Y-axis represents the absolute regression coefficient value. **e**) Utilizing each patients’ regression formula pre-vaccination and the known vaccination day, the RBD^+^ IgG level was extrapolated to predict the IgG level on the day of vaccination (i.e., day 0). Post-vaccination antibody levels were used to extrapolate the antibody levels on day 8 following vaccination. Y-axis represents the RBD^+^ antibody levels in log_10_. X-axis represents days in relation to the day of vaccination (i.e., day 0) at each serum collection time point and expected antibody levels. Dashed red lines indicate the upper and lower limit of negative control samples. **f**) Distribution of the recovered-vaccinated patients among quartiles as measured for their antibody levels at V1 compared to the extrapolated antibody levels at day 8 post-vaccination. Y-axis represents the percentage of recovered patients that were designated to each quartile. X-axis represents quartile groups. **g**) Comparison between the RBD^+^ IgG levels in recovered patients at V1, the extrapolated RBD^+^ IgG levels at day 0 (day of vaccine), day 8 following vaccination, and the RBD^+^ IgG levels of naïve vaccinees at an average of 8 DFV. Y-axis in log_10_. X-axis represents the relative days from vaccination of each sub-cohort. Dashed red lines indicate the upper and lower limit of the antibody levels in the negative control group.

## Discussion

The accelerated efforts to develop COVID-19 vaccines has borne fruit and following the release of the clinical trial data^25^, vaccine rollout campaign initiated globally utilizing the Pfizer-BioNTech BNT162b2 COVID-19 mRNA-vaccine (mRNA-vaccine)^26^. The mRNA-vaccine that is based on the coding region of the SARS-CoV-2 spike protein, is administered in two doses with an interval of 21 days between doses and to this end, approximately 60% of the Israel population received 2 vaccine doses. A study in a nationwide mass vaccination setting suggested that the mRNA-vaccine effectiveness (VE) is consistent with that of the randomized trial reported by Pfizer-BioNtech^27^. However, recently, the VE was reported to be decreased or impaired, and suggested to be associated with the emergence of the SARS-CoV-2 Delta variant of concern, and the possible waning of vaccine induced immunity ^28,29^. Priming the adaptive immune response by active vaccine, aims to protect against reinfection by the elicitation of vaccine-specific antibodies, memory B and T cells. The ideal vaccine should induce a robust immune response that is similar to the response following natural infection. This is the basic idea behind vaccine science – “training” the immune system towards a specific pathogen in a way that it will mimic the stimulation necessary for immune development, while not producing an active disease.

Highlighting the differences between the immune response following natural infection and vaccination may inform us whether the “training” provided by the vaccine, effectively stimulates the immune system which, in turn, will provide long-lasting immunity. Long-lasting immunity is an important part of the immune response and is attributed to the ability of the adaptive immune system to generate immune memory. The immune memory is composed of the serological and cellular arms (antibodies, memory T and B cells, respectively). Stable maintenance of antibody reservoirs can provide a mechanism to help mitigate subsequent infections. In the context COVID-19, the importance of the antibody reservoir is further highlighted by evidence of the temporal decay of the serological memory as observed in COVID-19 recovered patients and recent data suggesting that antibody levels wane in vaccinated individuals ^30,31^. Additionally, in a recent report by Israelow et al. ^32^ it was suggested that immune protection is largely mediated by antibody response and not cellular immunity highlighting the protective capacity of antibodies elicited by both natural infection and vaccination. Thus, it is of great importance to understand the longitudinal kinetics of antibodies which may provide long lasting protection. The longitudinal kinetics of RBD^+^ antibodies are the focus of this study aiming to evaluate the persistence of serological memory in COVID-19 recovered patients. The kinetics of RBD^+^ antibodies in COVID-19 recovered patients over 14 months reveal that following recovery, the patients experience decay in the antibody levels with IgA showing the fastest decay rate followed by IgM and IgG. This decay was observed by others, and it was suggested to be associated with the decreased reservoirs of long-lived plasma cells that are the source antibodies in circulation^33^.

However, the decay rate of antibodies in recovered patients is different between patients thus, it is important to examine the longitudinal kinetics at the individual level.

To do so, we examined the longitudinal kinetics of RBD^+^ antibodies in a cohort of 89 COVID-19 recovered patients. First, the patients were stratified to quartiles by the antibody levels they exhibit at the first visit (V1, mean 90 days DFS). Next, the RBD^+^ antibodies levels of sera samples from the patients of each quartile were used to calculate the regression coefficient that represents the decay rate. We found that the decay rate is affected by the robustness of the response as determined by the antibody levels at V1. Hence, patients that exhibit higher antibody levels at V1, experienced a faster decay rate. This decay trend was observed for all antibody isotypes.

Next, we aimed to understand the differences of antibody kinetics between recovered patients and naïve vaccinees who received 2 doses of mRNA-vaccine. Naïve vaccinees elicit a robust response and an average of 8 DFV, the IgG levels were designated to the highest quartile. However, the decay of all isotypes was found to be fast and at an average of 185 DFV, the antibody levels in 100% of individuals reached the threshold of the negative controls. Comparing the decay rate of IgG between COVID-19 recovered patients that exhibit the highest antibody levels (Q4) at V1 and the naïve vaccinees reveal a significantly faster decay in the naïve vaccinees group.

Overall, the data derived from this study provide new insights into the longitudinal kinetics of antibodies 14 months following recovery and highlights the difference of antibody persistence between recovered and vaccinees. Based on the results we hypothesize that the antibody longitudinal kinetics in recovered COVID-19 patients differs from the kinetics in naïve vaccinees due to fundamental differences between the mechanisms involved in the activation of the adaptive immune arm. While natural infection involves a full systemic activation including the innate immune arm, mRNA-vaccination may circumvent a full systemic activation. This circumference, in the case of mRNA-vaccine, could affect the ability of the immune system to maintain sufficient levels of long-lived plasma cells that supports the continuance of the antibody reservoir over time. In the case of COVID-19 recovered patients, it was demonstrated by Turner et al.^24^ that long-lived plasma cells persist over time, and this may explain the slower decay rate we detected in the recovered patient cohort. Noteworthy, these understandings were instrumental in facilitating the decision making in regards with the 3^rd^ vaccination regime strategy in Israel.

## EXPERIMENTAL MODEL AND SUBJECT DETAILS

### Human subjects

All patients and naïve individuals provided informed consent to the use of their data and clinical samples for the purposes of the study and blood collection was performed under institutional review board approvals number 0001281-4 and 0000406-1. Blood samples were collected at the Hasharon Hospital, Rabin Medical Center under ethical approval number 0265-20. A total of 345 sera samples from 200 recovered patients, all confirmed qPCR positive for SARS-CoV-2, were collected, with follow up samples from 89 COVID-19 recovered patients at approximately 90 days intervals at 3-4 visits (V1-V4). Additionally, 20 of the follow-up COVID-19 recovered patients received a single dose of BNT162b2 vaccine. Samples from 17 naïve individuals that received two doses of the BNT162b2 mRNA-vaccine were collected at four visits following mRNA-vaccine 2^nd^ dose as follows (median, days following vaccination - DFV): 8, 35, 91, and 182 DFV. Negative control included sera from 27 healthy individuals that were collected prior to the COVID-19 pandemic.

All blood samples were collected into BD vacutainer K2-EDTA collection tubes. Isolation of serum and peripheral blood mononuclear cells (PBMCs) was performed by density gradient centrifugation, using Uni-SepMAXI^+^ lymphocyte separation tubes (Novamed) according to the manufacturer’s protocol.

COVID-19 recovered participants were classified into levels of disease severity according to the classifying parameters given by the Israeli Ministry of Health (https://www.gov.il/en/departments/general/corona-confirmed-cases). Accordingly, patients that were confirmed for SARS-CoV-2 and exhibit fever, cough, malaise, and loss of taste and smell were considered as “mild”. Patients that were confirmed for SARS-CoV-2 and had pneumonia were considered as “moderate”. Patients that were confirmed for SARS-CoV-2 and had either respiratory frequency of more than 30 breaths per minute, or 93% saturation or less without supplements or problems in oxygenation was considered “severe”. Any case that showed more severe parameters, such as the requirement of ventilation and/or suffers from multiple organ dysfunction, will also be in the “severe” category for our study. COVID-19 recovered participants were stratified into two groups: one consists of all “mild” cases and another of the “moderate” and “severe” cases.

## Method Details

### Expression and purification of recombinant protein

The plasmids for expression of recombinant SARS-CoV-2 receptor-binding domain (RBD) was kindly provided by Dr. Florian Krammer, Department of Microbiology, Icahn School of Medicine at Mount Sinai, New York, NY, USA. The RBD sequence is based on the genomic sequence of the first virus isolate, Wuhan-Hu-1, which was released on January 10th, 2020, ^9^. The plasmids for expression of recombinant human ACE2 (hACE2) was kindly provided by Dr. Ronit Rosenfeld from the Israel Institute for Biological Research (IIBR). The cloned region encodes amino acids 1-740 of hACE2 followed by 8xHis tag and a Strep Tag at the 3’ end, cloned in a pCDNA3.1 backbone. Recombinant RBD and hACE2 were produced in Expi293F cells (Thermo Fisher Scientific) by transfections of these cells with purified mammalian expression vector using an ExpiFectamine 293 Transfection Kit (Thermo Fisher Scientific), according to the manufacturer’s protocol, and as described previously ^9^. Supernatants from transfected cells were purified on HisTrap affinity column (GE Healthcare) using 2-step elution protocol with 5CV of elution buffer supplemented with 50mM imidazole in PBS, pH 7.4 followed by 250mM imidazole in PBS, pH 7.4. Elution fractions containing clean recombinant proteins were merged and dialyzed using Amicon Ultra (Mercury) cutoff 10K against PBS (pH 7.4). Dialysis products were analyzed by 12% SDS–PAGE for purity and concentration was measured using Take-5 (BioTek Instruments).

Purified recombinant proteins were biotinylated using the EZ-Link Micro-NHS-PEG4-Biotinylation kit (Thermo Scientific), according to the manufacturer’s protocol.

### Serum anti-SARS-CoV-2 RBD antibodies assay

RBD^+^ Ig levels in sera were determined using 96 well ELISA plates that were coated overnight at 4°C with 2μg/ml RBD in PBS (pH 7.4). Next, coating solution was discarded, and ELISA plates were blocked with 300µl of 3% w/v skim milk in PBS for 1 hour at 37°C. Following the discarding of the blocking solution, duplicates of serum diluted 1:300 in 3% w/v skim milk in PBS were added to the microwells. Negative control serum samples were also added in single wells of 1:300 dilutions, with a range of 8-24 control in each plate. ELISA plates were then washed three times with PBST and 50μl of horseradish peroxidase (HRP) conjugated goat anti-human IgG (Jackson ImmunoResearch, #109035003) / anti-human IgM (Jackson ImmunoResearch, #109035129) / anti-human IgA (Jackson ImmunoResearch, #109035011) secondary antibodies were added to each plate at the detection phase (50μl, 1:5000 ratio in 3% w/v skim milk in PBS) and incubated for 1 hour at room temperature (RT), followed by three washing cycles with 0.05% PBST. Development was carried out by adding 50µl of TMB for 5 minutes of exposure, immediately followed with reaction quenching by adding 0.1M sulfuric acid. Plates were read using the Epoch Microplate Spectrophotometer ELISA plate reader using wavelengths of 450 and 620 nm. Reads from the 620 nm wavelength were reduced from those received by the 450 nm wavelength, resulting in a final signal value for each well, and therefore 2 signal duplicates for each tested serum and one for each negative control serum. A total “Negative control value” for each plate was computed as the average of all negative control signals, in addition to one standard division value. A “signal over negative” (S/N) value for each well was then produced. All sera were evaluated in a second independent technical repeat, resulting in 4 S/N values for each serum, that were averaged to a single RBD^+^ Ig titer level.

### Computational and statistical analysis

#### Bootstrapping & quartile establishment

We stratified the RBD^+^ antibody levels to quartiles by musing the bootstrap method. Using a R code script, 50 randomly selected values from the extended sera sample cohort (n=345) were used to set quartile thresholds according to the mean values derived from the bootstrap iterations, a process which was then repeated 10,000 times. The produced 10,000 quartile values were then averaged to 4 final quartile threshold values (Figure S1). These values were then used to designate each measured antibody level at V1 to its respective quartile range (i.e., from low to high Q1-Q4 respectively). The benefit of bootstrapping data is in its ability to achieve minimal CI inference and minimize the effects of outlier data points in relatively small sample sizes that might not be evenly distributed ^34^.

#### Statistical analysis

Scatter plot regression was analyzed using a nonlinear regression model (Fig. 2b, Fig. 3b, Fig. 4a, Fig 4c). Correlations between titer results of ELISA technical repeats were analyzed using Spearman’s rank correlation. A Mann-Whitney test was used to compare two independent groups with continuous variables (Fig. 2a and Fig. 4g). A Comparison of fits test was used to compare nonlinear regression models (Fig. 2c., Fig. 3c., Fig. 4b and Fig. 4d). A Wilcoxon test was used to compare two or more sets of dependent groups Fig. 4g.). All reported P values were two-tailed, and p values less than 0.05 were considered statistically significant. All statistics were performed with GraphPad Prism 9.0.2 (GraphPad Software).

## Data Availability

The data that support the findings of this study are available on request from the corresponding author [D.D. or W.Y.]. The data are not publicly available due to the fact that they contain information that could compromise research participant privacy/consent.

## Author Contributions

E.T., V.M.A., T.K.R., M.D., D.D and W.Y initiated and designed the study, T.K.R., K.T, M.D. and D.D. recruited study participants and preformed blood collections, E.T., V.M.A., D.Y., A.L., K.A., R.S., B.Y. and T.D performed blood separation, E.T., V.M.A performed ELISA assays, E.T., A.L., T.D., and W.Y. performed statistical analysis, E.T., W.Y and D.D wrote the manuscript. All authors reviewed and approved the manuscript.

## Acknowledgment

We wish to thank Prof. Itai Benhar and Dr. Limor Nahary from the Shmunis school of Biomedicine and cancer research, Tel Aviv University, for their assistance in providing the initial RBD and hACE2 reagents, Dr. Ronit Rosenfeld from the Israel Institute for Biological Research (IIBR) for her generous assistance and Prof. Mordechai Gerlic and Prof. Ariel Munitz from the Sackler Faculty of medical, Tel Aviv University. This work was supported by the Israel Ministry of Health (MOH) grant #3-17162.

