## supplementary information for "The longitudinal kinetics of antibodies in COVID-19 recovered patients over 14 months"

**a.**

| | | | | | |
|---|---|---|---|---|---|
| Total COVID-19 Recovered patient cohort | Age | Max | 81 | Average | 53.5 |
| | | Min | 20 | | |
| | Gender | M | 38 | Total | 82 |
| | | F | 44 | | |
| F | Age | Max | 78 | Average | 53.8 |
| | | Min | 20 | | |
| M | Age | Max | 81 | Average | 53 |
| | | Min | 25 | | |

**b.**

| | | | | | |
|---|---|---|---|---|---|
| Follow up COVID-19 Recovered patient cohort | Age | Max | 81 | Average | 50.5 |
| | | Min | 20 | | |
| | Gender | M | 47 | Total | 90 |
| | | F | 43 | | |
| F | Age | Max | 78 | Average | 49 |
| | | Min | 20 | | |
| M | Age | Max | 81 | Average | 53 |
| | | Min | 25 | | |

**c.**

| | | | | | |
|---|---|---|---|---|---|
| Follow up COVID-19 Recovered patient cohort who received one BNT162b2 mRNA-vaccine dose | Age | Max | 76 | Average | 48 |
| | | Min | 20 | | |
| | Gender | M | 13 | Total | 22 |
| | | F | 9 | | |
| F | Age | Max | 76 | Average | 48 |
| | | Min | 20 | | |
| M | Age | Max | 76 | Average | 52 |
| | | Min | 28 | | |

**d.**

| | | | | | |
|---|---|---|---|---|---|
| Naïve vaccinees | Age | Max | 65 | Average | 43.5 |
| | | Min | 22 | | |
| | Gender | M | 4 | Total | 17 |
| | | F | 13 | | |
| F | Age | Max | 65 | Average | 43.5 |
| | | Min | 22 | | |
| M | Age | Max | 63 | Average | 46 |
| | | Min | 29 | | |

**Table S1 | Study participant cohorts a)** COVID-19 recovered patients **b)** Follow up recovered patients (sub-cohort of the recovered patients). **c)** Follow up recovered patients that received one BNT162b2 mRNA-vaccine dose. **d)** Naïve vaccinees that received two doses of the BNT162b2 mRNA-vaccine.

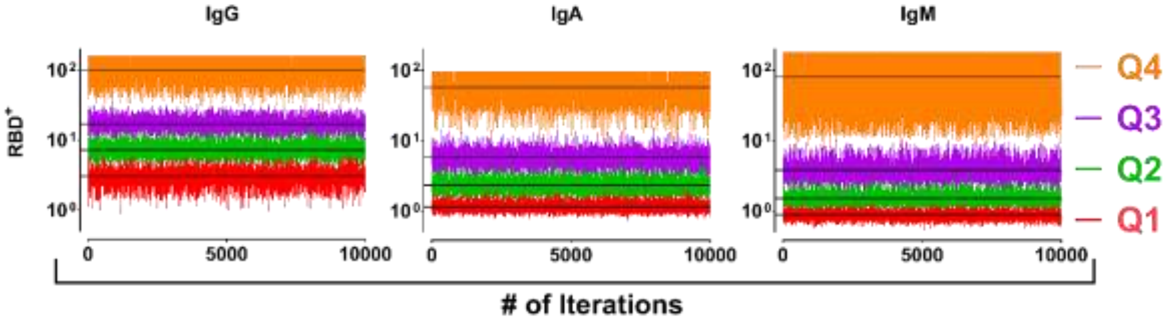


**Figure. S1 | Quartile determination utilizing bootstrap method.** RBD$^+$ antibody measured values were randomly selected (n=50) out of the extended dataset obtained from all recovered patients (n=345) and quartile thresholds were set according to the mean values derived from 10,000 iterations. Y-Axis in $Log_{10}$, X-axis represent the number of iterations used in the bootstrapping procedure.

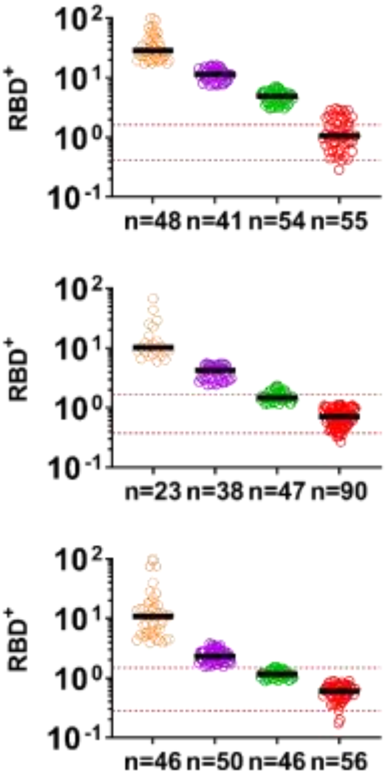


**Figure. S2 | Designation of RBD$^+$ antibody levels by quartiles.** Measured RBD$^+$ antibody levels in recovered patients were designated to their respective quartile (i.e., from low to high Q1-Q4 respectively) according to the measured antibody level at the first visit (V1). Y-axis, $Log_{10}$ of RBD$^+$ antibody levels. X-axis the quartile groups, n indicates the number of individuals assigned to each quartile at V1 for each isotype.
